# Neurohistopathological Findings of the Brain Parenchyma After Long-Term Deep Brain Stimulation: Case Series and Systematic Literature Review

**DOI:** 10.1101/2023.11.23.23298888

**Authors:** Juan Vivanco-Suarez, Timothy Woodiwiss, Kimberly L. Fiock, Marco M. Hefti, Nandakumar S. Narayanan, Jeremy D. W. Greenlee

**Author notes:** Both authors contributed equally. **CORRESPONDENCE** Jeremy D. W. Greenlee, MD, Department of Neurosurgery, University of Iowa Hospitals and Clinics, 200 Hawkins Dr, 1826 JPP, Iowa City, IA 52242, United States. E-mail address. **PREVIOUS PRESENTATION**. **ETHICS**. **CONFLICTS OF INTEREST**. The authors report no disclosures. **FUNDING**. This research received no specific grant from any funding agency in public, commercial, or not-for-profit sectors.

## Abstract

**Introduction:** The efficacy of deep brain stimulation (DBS) has been established to treat several movement and psychiatric disorders. However, several aspects, such as the mechanism of action and tissue changes after treatment are incompletely described. Hence, we aimed to describe the neurohistopathological findings of 9 patients who underwent DBS for movement disorders. Additionally, we performed a systematic literature review on postmortem studies after DBS implantation.

**Methods:** We performed a retrospective study of patients who underwent DBS for movement disorders between 2000-2023 and had postmortem autopsy. Demographics, clinical features, and outcomes were collected. Levodopa equivalent daily dose (LEDD) and total electrical energy delivered (TEED) were calculated. Neurohistopathological assessments were performed from autopsies. A systematic literature review was conducted to summarize the literature.

**Results:** Postmortem neurohistopathologic assessment of 9 patients who underwent DBS for movement disorders (8 Parkinson’s disease [multiple system atrophy was final diagnosis in 1], and 1 parkinsonism) was performed. Median age at DBS implantation was 65 years (range, 54-69), and most were male (8 patients, 89%). Most common DBS target was the subthalamic nucleus (8 patients, 89%). Median DBS duration was 65 months (range, 7-264 months). Post-DBS reduction in LEDD was found in 7/9 patients, and TEED was increased over time in 5/7 patients. No patients died due to DBS. Neurohistopathological assessment showed gliosis in 7 (78%) patients and activated microglial infiltration in 1 (11%). Additionally, postmortem findings (after DBS) of 59 patients (between 1977-2021) were identified in the literature: 26 (44%) for Parkinson’s disease, 20 (34%) for pain, and 13 (22%) for other conditions.

**Conclusion:** Our findings confirm the presence of a local tissue reaction, including gliosis and activated microglial infiltration around the implanted DBS electrodes. The effect of the local changes on the clinical efficacy of DBS is not established. Further DBS postmortem studies and standardization of tissue processing are needed.

## INTRODUCTION

The efficacy of deep brain stimulation (DBS) has been established for the treatment of several movement and psychiatric disorders.^1^ Furthermore, its experimental application for chronic pain, obesity, and other neurocognitive disorders has continually expanded, aiming to benefit a broader group of patients.^2^ However, several aspects of this treatment are not completely defined, including the specific interaction and activation of surrounding structures, the local tissue changes in the brain caused by long-term stimulation, the potential adverse tissue responses induced by the electrode, and the existence of neuronal damage.^1,3^ A deeper understanding of the chronic local glial responses after electrode implantation may provide helpful insights for the improvement of anatomical targets and the optimization of electrode design according to the target tissue.

Previously published postmortem studies have begun the characterization of histological changes occurring around electrodes in response to implantation and chronic stimulation. A comprehensive review performed by DiLorenzo et al. reported 40 cases with implanted systems from as early as 1977.^4^ The authors showed that in more than 50% of the cases, there was local fibrosis, astrocytosis, multinucleated giant cells, mononuclear leukocytes, macrophages, activated microglia, and neuronal loss.^4,5^ Of note, persistent efficacy was more commonly observed when tissue injury was absent. More recently, Vedam-Mai et al. found that 3 out of 4 DBS cases exhibited histopathological evidence of a glial collar or scar around the electrode, but there was no significant association between DBS duration and the amount of gliosis. As the technologies and techniques employed for DBS continue to evolve, the evidence regarding the local changes around the electrode is contradictory.^6–9^ For instance, data on the comparison of histological samples from patients with active and inactive electrodes is limited, the effect of the size of the gliotic scar is uncertain, and evidence of the distance (from the electrode) where neuronal loss or preservation is observed is unclear.^10^

Hence, this study aims to characterize and describe the neurohistopathological environment of the brain parenchyma surrounding DBS electrodes in 9 patients who underwent surgical treatment for Parkinson’s disease (PD) or a parkinsonism syndrome. Additionally, we present an updated and expanded summary of the literature on this topic with a systematic review of postmortem studies after DBS implantation.

## METHODS

### Study design, patient protocol, tissue acquisition, and storage

This was a retrospective study of adult patients who underwent DBS for movement disorders at a tertiary academic center between July 2000 and April 2023. All tissue was obtained from autopsies conducted at the University of Iowa Hospitals and Clinics as part of routine clinical care in accordance with all applicable federal and state laws and regulations. Consent for the use of tissue in research was obtained from the next of kin as part of the autopsy consent. This project was reviewed by the University of Iowa Institutional Review Board and determined not to represent human subjects research under the National Institutes of Health Revised Common Rule. Based on the database created for each donor, for this study, we extracted the age at diagnosis, sex, duration of disease, calculated Levodopa equivalent daily dose (LEDD)^11^ (before implantation, 6 [± 3] months after DBS, and at last clinical follow-up), age at DBS implantation, intraoperative and hospitalization complications, DBS duration, post-implantation stimulation parameters (at 6 [± 3] months after DBS, at last clinical follow-up), and age at death. To assess the stimulation parameters, we calculated total electrical energy delivered (TEED)^12^ (impedance was set to 1000Ω to standardize calculations).

### Neurohistopathological techniques and evaluation

Tissue processing was done as previously described.^13–15^ Briefly, brains were fixed in 20% neutral buffered formalin for 10-14 days and then sectioned, sampled, processed, and stained (both histochemical and immunohistochemical stains) in the Emory Warner Clinical Pathology Laboratories of the University of Iowa according to standard protocols. A board-certified neuropathologist (MMH) and PhD-trained research neuropathologist (KLF) reviewed all tissue sections and stains.

### Systematic literature review and data extraction

A comprehensive literature search of the MEDLINE (via PubMed) database using medical subject heading terms, entrees, and free text to identify all published postmortem neurohistopathological reports of DBS cases (end-of-search date: August 9, 2023). The complete search strategy is found in **Supplemental Table 1**. All DBS cases, regardless of indication or anatomical target in the human brain, were included. Review articles or meta-analyses that did not describe disaggregated data on DBS cases were excluded. Two independent reviewers (TW and JV-S) screened the articles using the Rayyan tool (https://www.rayyan.ai/). Any difference between reviewers was settled through discussion. After the initial selection of eligible studies, the reference lists were thoroughly searched to identify additional eligible studies based on the “snowball” methodology.^16^ Because DiLorenzo et al. thoroughly reviewed the literature in 2014, we expanded our review to new cases either missed by the aforementioned authors or reported after the review was completed. The following data were collected using a standardized data extraction form: demographic and patient characteristics, information about implanted hardware, stimulation parameters, clinical course, and neuropathological findings. Clinical efficacy and decline after DBS were extracted as defined by the authors of each study. When critical or disaggregated information was missing, the studies were excluded.

## RESULTS

### Patient demographics, clinical, and DBS characteristics

A total of 9 patients who had DBS implantation were identified from our institutional autopsy database. The patient and neurohistopathologic characteristics are described in **Table 1**. The median age of disease onset was 52 (range, 47-64 years), and 8 (89%) were male. On preoperative clinical evaluation, PD was diagnosed in 8 (89%) patients and parkinsonism in 1 (11%). Of note, patient 9 presented clinically as PD with a very robust response to levodopa for several years. However, dysautonomic symptoms and ataxia emerged throughout the disease course, pointing to the diagnosis of a slowly evolving Parkinsonian variant multiple system atrophy (MSA), which was confirmed on postmortem neurohistopathologic evaluation.

**Table 1.**
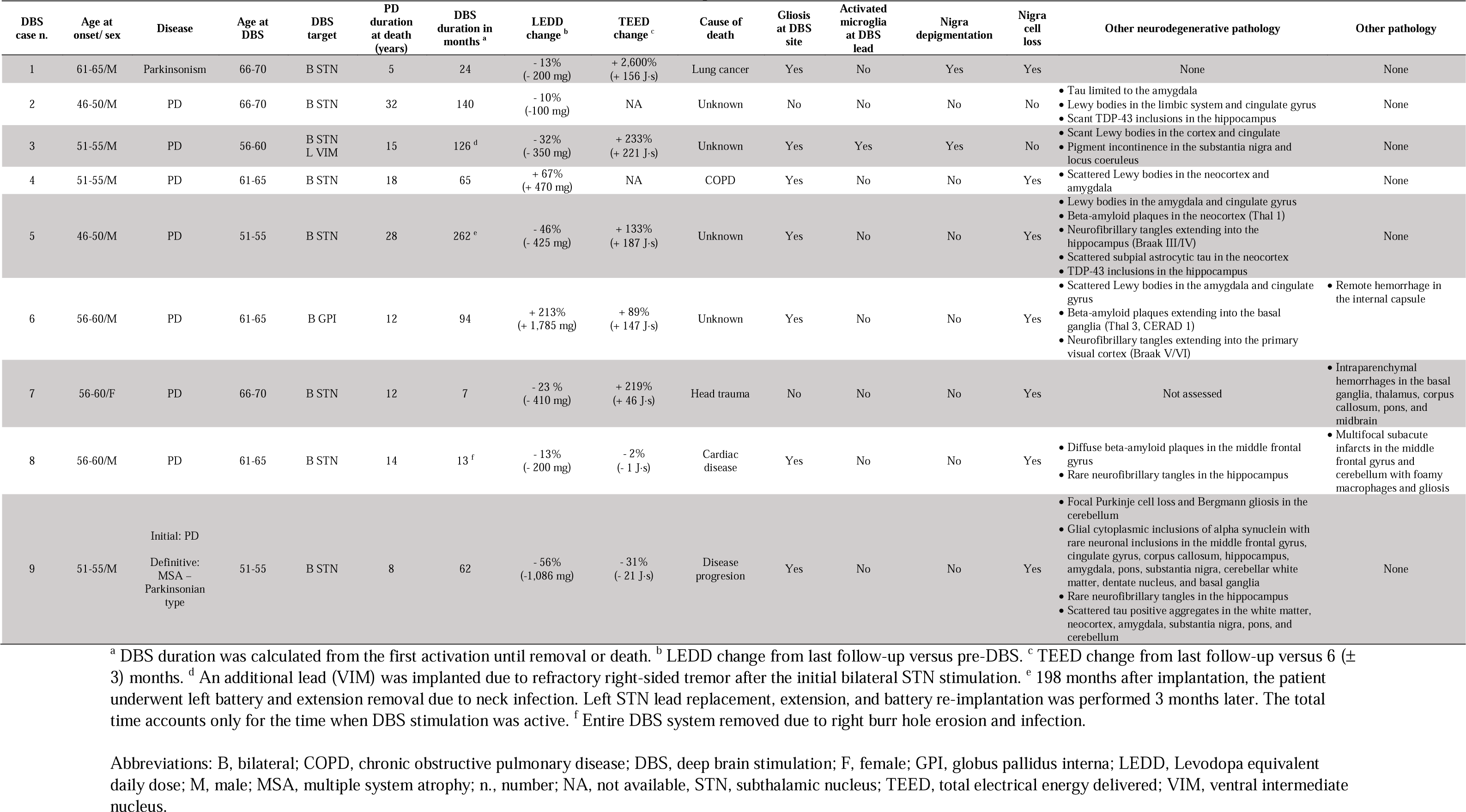
Demographic, clinical, and neurohistopathological (around the electrode track) characteristics of the patients who underwent DBS implantation.

Regarding the DBS characteristics, the median disease course before DBS implantation was 7 years (range, 3-20 years). The median patient age at DBS implantation was 65 years (range, 54-69 years). All patients received bilateral DBS lead implantation; the majority received electrodes targeting the bilateral subthalamic nuclei (8, 89%). One (11%) patient received leads targeting the globus pallidus interna. Patient 3 had an additional lead implanted in the left ventralis intermedius thalamic nucleus due to refractory right-sided tremor. All electrodes were made of platinum/iridium (Medtronic, Inc.). The median DBS duration was 65 months (range, 7-262 months). Patient 5 underwent unilateral DBS removal (extension and battery) 16 years after bilateral implantation due to a neck infection surrounding the left-sided extension wire. No intracranial infection was identified. He was successfully reimplanted 3 months later after adequate infection management with replacement of the left STN lead and extension and battery replacement. Patient 8 had a right lead incision/burr hole erosion and infection necessitating complete bilateral DBS hardware removal 13 months after implantation. No intracranial infection was identified At last follow-up, the TEED increased in 5 of 7 patients with available data by a mean of 151 J·s compared to their initial DBS settings. However, in patients 8 and 9, the TEED decreased slightly (1 and 21 J·s, respectively) from their initial parameters. A mean LEDD reduction of 28% was observed in 7 patients after DBS began. On the other hand, LEDD increased in patients 4 and 6 by 67% and >100%, respectively. Additional details about the LEDD and TEED are presented in **Supplemental Table 2**. In terms of mortality, 1 patient (11%, case 9) passed away due to disease progression, 4 (44%) died due to unknown causes, and the remaining 4 (44%) passed away due to reasons unrelated to the primary disease or an unknown cause.

### Postmortem neurohistopathological characteristics

An example of the gross and microscopic findings is presented in **Figure 1 (A-E)**. **Table 1** shows a descriptive summary of the postmortem neurohistopathologic findings. All patients except 2 showed gliosis at the DBS lead site. One of the 2 only had DBS for 7 months due to a fatal fall, while the other showed no gliosis despite DBS for 140 months. Activated microglial infiltration was seen around the lead in 1 patient. For the 7 patients who developed gliosis around the DBS lead, the median DBS duration was 65 months (range, 13-262). TEED increased over time in 4 patients (mean of 178 J·s) and decreased in 2 (1 and 21 J·s, respectively). A mean LEDD reduction of 32% was observed in 5/7 patients after DBS compared to pre-operative values. Two patients required an increase of their LEDD over time of 67% and >100%, including the lone PD patient treated with globus pallidus interna DBS. Notable neurohistopathologic changes from the tissue sections are the following: substantia nigra depigmentation was found in 2 (22%) patients (cases: 1 and 3) and nigra cell loss in 7 (78%; cases: 1, 4, 5, 6, 7, 8, and 9). Patient 1 had a combination of nigra depigmentation and nigra cell loss. There were a variety of additional neurodegenerative pathologic findings, including the presence of Lewy bodies in 5 patients (56%; cases: 2, 3, 4, 5, and 6) at different anatomical structures, neurofibrillary tangles in 4 (44%; cases: 5, 6, 8, and 9), tau aggregates in 4 (44%; cases: 2, 5, 8, and 9), beta-amyloid plaques in 3 (33%; cases: 5, 6, and 8), TDP-43 inclusions in 2 (22%; cases: 2 and 5), and inclusions of alpha-synuclein in 1 (11%, case 9). Of note, all but one assessed patients with 12 years or longer of PD duration at the time of death showed Lewy bodies. Finally, other pathological findings (distant to the tract) including hemorrhages and subacute infarcts were reported in 3 (33%) patients (cases 6, 7, and 8).

**Figure 1(A-E).**
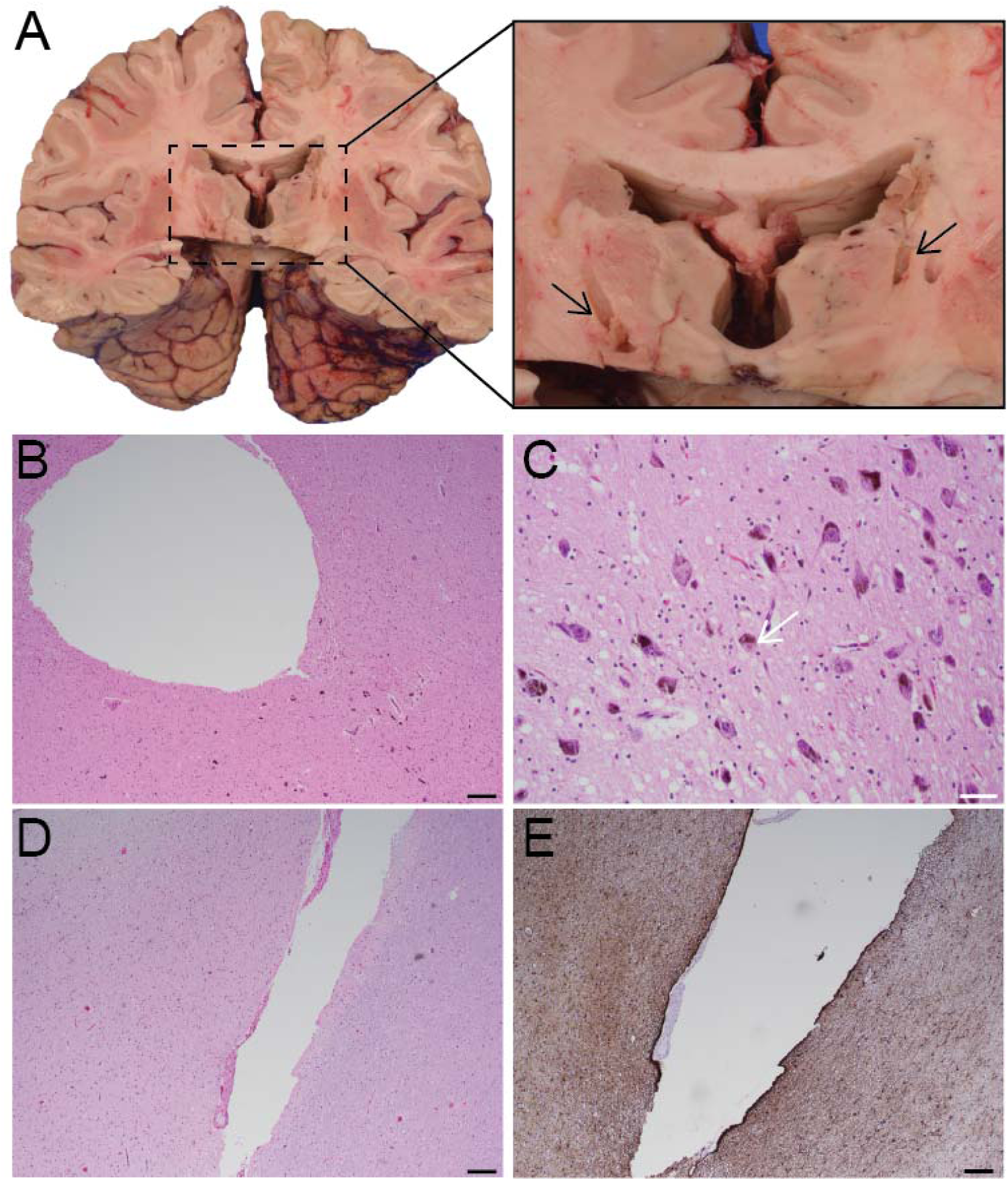
Macroscopic and microscopic tissue representations. (**A**) Coronal brain slice depicting gross bilateral tracts with close-up pop-out. Arrows denote tracts. (**B**) Hematoxylin & Eosin stain slide of midbrain with electrode tract in the nigra. Black scale = 200 µm. (**C**) Lewy bodies in the nigra, arrow denotes Lewy body. White scale = 50 µm. (**D**) Luxol fast blue stain on thalamus with electrode tract. Black scale = 200 µm. (**E**) Glial fibrillary acidic protein statin on thalamus with tract. Black scale = 200 µm.

### Systematic review update: study selection and characteristics

The literature review yielded 1249 articles, from which 18 duplicates were removed. As a result of the initial screening by title and abstract, there were 48 potentially eligible articles. In the full-text evaluation, 16 documents were excluded for the following reasons: ineligible study type (*n*=6), insufficient data available (*n*=5), ineligible outcomes (*n*=3), and ineligible patient population (*n*=2). Ultimately, 32 studies (25 case reports and 7 case series) from the final systematic search were included.

From the studies, a total of 59 patients who had a postmortem neurohistopathological assessment after DBS implantation (for different conditions) were identified. The studies were performed between 1977 and 2021. Fifteen were conducted in Europe ^6,7,10,17–28^, 13 in North America^29–41^, 3 in Australia^42–44^, and 1 in Asia^45^. The number of patients per study ranged from 1 to 8. In terms of the conditions for which DBS was implanted, 26 (44%) patients were treated for PD between 1994 and 2021 (**Table 2**; 2 MSA cases were initially misdiagnosed as PD), 6 (10%) for epilepsy (4 cases had cerebellar stimulation) between 1979 and 2020 (**Supplemental material Table 3**), 20 (34%) for pain between 1977 and 1991 (**Supplemental material Table 4**), and 7 (12%) for other conditions (tremor, chorea-acanthocytosis, and diffuse Lewy body disease) between 2000 and 2011 (**Supplemental material Table 5**).

**Table 2.** Characteristics of the postmortem neurohistopathological findings around the DBS electrode track for Parkinson’s disease.

| Author, year | DBS case n. | Age at DBS | Sex | DBS targets | Electrode surface material | Clinical efficacy after DBS | Clinical decline after DBS | DBS duration in months | Cause of death | Gliosis at DBS site | Activated microglia at DBS lead | Neuronal loss at DBS site | Other findings at DBS site | Nigra depigmentation | Nigra cell loss |
| --- | --- | --- | --- | --- | --- | --- | --- | --- | --- | --- | --- | --- | --- | --- | --- |
| Caparros-Lefebvre et al., 1994<sup>17</sup> | 1 | 56-60 | M | Vim | Pt/Ir | Yes | No | 43 | Stroke | Yes (1 mm perimeter) | NR | NR | Accumulation of lymphoid cells, macrophages, and giant cells (2 mm) around the electrode | Yes | Yes |
| Haberler et al., 2000<sup>10</sup> | 2 | NR | NR | L Vim | Pt/Ir | Yes | No | 70 | Myocardial infarction | Yes | No | No | - | Yes | Yes |
| “ | 3 | 71-75 | M | BL Vim | Pt/Ir | Yes | No | 54 | Cardiovascular failure | Yes | No | No | Mononuclear leukocytes | Yes | Yes |
| “ | 4 | 51-55 | M | L Vim | Pt/Ir | Yes | No | 45 | Cardiovascular failure | Yes | No | No | Macrophages and reactive astrocytes | Yes | Yes |
| “ | 5 | 56-60 | M | BL Vim | Pt/Ir | Yes | No | 45 | Cardiovascular failure | Yes | NR | No | Multinucleated giant cells and reactive astrocytes | Yes | Yes |
| “ | 6 | 41-45 | M | R Vim | Pt/Ir | Yes | No | 1 | Astrocytoma | Yes | NR | No | Multinucleated giant cells | Yes | Yes |
| “ | 7 | 76-80 | M | BL Vim | Pt/Ir | Yes | No | 15 | Cardiovascular failure | Yes | Yes | No | More pronounced gliosis around lead track, multinucleated giant cells, and reactive astrocytes | Yes | Yes |
| “ | 8 | 66-70 | F | BL STN | Pt/Ir | Yes | No | 3 | Drowning | Yes | Yes | No | More pronounced gliosis around lead track, multinucleated giant cells, and reactive astrocytes | Yes | Yes |
| “ | 9 | 61-65 | M | L STN | Pt/Ir | Yes | No | 2 days | Myocardial infarction | - | No | No | Fresh hemorrhage, perifocal edema, and numerous axonal spheroids | Yes | Yes |
| Henderson et al., 2001<sup>42</sup> | 10 | 66-70 | M | L CM-Pf | Pt/Ir | Yes | Yes | 48 | Sepsis | Yes | NR | Yes | Extensive astrocytosis and small thalamotomy greater than the dimension of the electrode tip penetration | Yes | Yes |
| Berciano et al., 2002<sup>18</sup> | 11* | 61-65 | M | BL STN | NR | Yes | No | 21 days | Pulmonary embolus | NR | NR | NR | NR | Yes | Yes |
| Henderson et al., 2002<sup>43</sup> | 12 | 51-55 | M | BL STN | Pt/Ir | Yes | No | 2 | Aspiration pneumonia | Yes | Yes | Yes (mild) | TUNEL staining and analysis of cellular morphology did not show features of apoptosis, and astrocytosis | Yes | Yes |
| Counelis et al., 2003<sup>29</sup> | 13 | 61-65 | M | BL STN | Pt/Ir | Yes | - | 4 days | Pulmonary embolus | No | No | No | Fresh hemorrhage along tracks and minimal reactive changes | NR | Yes |
| Jarraya et al., 2003<sup>19</sup> | 14 | 21-25 | F | BL STN | NR | Yes | Yes | 24 | Pulmonary embolus | Yes | NR | NR | Large number of activated macrophages around the cavity and in the vicinity, and astrocytosis | Yes | Yes |
| Chou et al., 2004<sup>30</sup> | 15* | 51-55 | M | BL STN | NR | No | Yes | 3 | Aspiration pneumonia | Yes | NR | NR | Tracks associated with hemosiderin deposition, multinucleated giant cells, and neovascularization | Yes | Yes |
| Henderson et al., 2004<sup>44</sup> | 16 | 61-65 | M | L Vim | Pt/Ir | Yes | Yes | 84 | Ischemic heart disease | Yes | No | Yes | Hemosiderin deposition along the electrode track and astrocytosis | Yes | Yes |
| Nielsen et al., 2007<sup>20</sup> | 17 | 71-75 | M | BL STN | Pt/Ir | Yes | No | 29 | Accident | Yes (capsule) | NR | No | Lymphocytes, macrophages, neutrophilic granulocytes, occasional neurons in the capsule, and astrocytosis | Yes | Yes |
| Guehl et al., 2008<sup>21</sup> | 18 | 51-55 | M | BL ZI | Pt/Ir | Yes | No | 72 | Dystonia (rhabdomyolysis) | Yes | NR | NR | None of the electrode contacts were located within the STN | NR | NR |
| Sun et al., 2008<sup>31</sup> | 19 | 81-85 | M | BL STN | Pt/Ir | Yes | No | 71 | Pneumonia | Yes | Yes | NR | Capsule with hemosiderin-laden macrophages, scattered lymphocytes, Rosenthal fibers, focal foreign-body giant cell reaction, and astrocytosis | Yes | Yes |
| Vedam-Mai et al., 2012<sup>32</sup> | 20 | 71-75 | M | BL STN | NR | Yes | No | 13 | Cardiac arrest | Yes | No | No | Marked fibrosis around the cavity (right DBS lead tip), fibrocollagenous “capsule” confirmed by Mason’s trichrome | Yes | Yes |
| Al-Helli et al., 2015<sup>6</sup> | 21 | 66-70 | M | BL STN | NR | NR | NR | 72 | NR | Yes | Yes | NR | Small collagenous scar with gliosis (Perl’s stain positive) around track | Yes | Yes |
| De Vloo et al., 2018<sup>7</sup> | 22 | 66-70 | F | BL STN | Pt/Ir | Yes | No | 144 | Pneumonia | Yes | No | No | Minimal reactive astrocytosis, macrophages, fibrillary gliosis, and perivascular lymphocytic infiltration | Yes | Yes |
| Gelpi et al., 2021<sup>22</sup> | 23 | 61-65 | M | BL STN | NR | Yes | No | 51 | Peritonitis | Yes | Yes | Yes | Astrogliosis | Yes | NR |
| “ | 24 | 66-70 | F | BL STN | NR | Yes | No | 52 | Pneumonia | Yes | Yes | Yes | Rosenthal fibers and astrogliosis | Yes | NR |
| “ | 25 | 46-50 | M | BL STN | NR | Yes | No | 36 | Pneumonia | Yes | Yes | Yes | Multinucleated giant cells, Rosenthal fibers, and astrogliosis | Yes | NR |
| “ | 26 | 51-55 | M | BL STN | NR | Yes | No | 48 | Cardiac arrest | Yes | Yes | Yes | Rosenthal fibers and astrogliosis | Yes | NR |

\* DBS indicated for PD, but postmortem examination established diagnosis of multiple system atrophy Abbreviations: BL, bilateral; CM-Pf, centromedian-parafascicular complex thalamic region; DBS, deep brain stimulation; F, female; L, left; M, male; n., number; NR, not reported; Pt/Ir, platinum/iridium; R, right; STN, subthalamic nucleus; TUNEL, (TdT)-mediated UTP nick and labelling; Vim, ventralis intermedius thalamic nucleus; ZI, zona incerta.

### Literature PD patients: DBS characteristics

Regarding the 26 PD patients (cases 11^18^ and 15^30^ were confirmed to be MSA on autopsy), the age at DBS implantation ranged from 24 to 81 years, and most were males (21; 81%). The DBS targets were the subthalamic nucleus in 16 (62%) patients (15 bilateral and 1 left side), the ventralis intermedius thalamic nucleus in 8 (31%; 3 bilateral, 3 right, 2 left, and 1 unknown), the left centromedian-parafasicular complex thalamic region in 1 (4%), and the bilateral zona incerta in 1 (4%). Platinum/iridium electrodes were the most employed. DBS duration ranged from 2 days to 144 months. Three cases passed away within 1 month after DBS implantation. Case 9 passed away 2 days after implantation due to myocardial infarction.^10^ Case 11 was readmitted 19 days after lead placement surgery due to abnormal behavior, impaired gait, fever, and lethargy. DBS was discontinued, and medical management was started.^18^ However, after transient improvement, the patient died 2 days later due to a massive pulmonary embolism (identified in autopsy). Case 13 died 4 days after implantation due to cardiorespiratory arrest secondary to pulmonary embolus (identified in autopsy). Clinical efficacy (as reported by the authors of each study) was observed in 24 patients (92%), and decline was reported in 4 (15%). Death causes were diverse. However, none of them were encountered intraoperatively during DBS implantation.

### Literature PD patients: postmortem neurohistopathological characteristics

In terms of the postmortem neurohistopathological analysis, fibrillary gliosis (which represents a chronic tissue reaction) was reported in 23/24 (92%) patients, and in 2/26 (8%) patients, it could not be assessed due to precipitated death (details discussed above). In the patients who passed away within 1 month after implantation, fresh hemorrhage was evidenced in 2/3 (67%). Reactive astrocytosis (mostly reflective of an acute tissue reaction) was observed in 14/25 (56%) patients. Microglial activation was found in 9/17 (53%) patients, and neuronal loss was observed in 7/19 (37%) patients. Multinucleated giant cells were reported in 8/25 (32%) patients. Regarding the pathologic findings of PD, nigra depigmentation was reported in 24/24 (100%) patients, and nigra cell loss in 21 (81%). Of note, in the cases where MSA was confirmed on autopsy (2/26, 8%), DBS showed clinical efficacy only in case 11.

## DISCUSSION

In this study of the neurohistopathological findings of 9 patients implanted with platinum/iridium DBS electrodes for parkinsonism (1 patient), PD (7 patients), and parkinsonian type MSA (1 patient), we found that at a median time of 65 months after DBS implantation, the brain tissue surrounding the DBS resulted in gliosis in 7 patients (78%) and the presence of activated microglia in 1 patient (11%). In general, these results are consistent with previous postmortem studies showing that a glial response is consistently generated around the DBS electrodes. Additionally, our systematic review spanning between 1977 and 2021 of postmortem neuropathological studies reporting findings after DBS identified 59 unique patients who underwent treatment for PD, epilepsy, pain, and other conditions such as tremor, chorea-acanthocytosis, and diffuse Lewy body disease. As the number of patients treated with DBS (for a wide variety of conditions) increases and the technologies continue to evolve, our study represents an addition to the body of evidence on the postmortem findings after DBS and represents the most comprehensive and current review of this topic.

### Gliosis surrounding the implantation site

The presence of a gliotic response surrounding the DBS electrode described in this study is similar to the findings reported in previous postmortem human studies and animal models.^2^ Yet, our rate of 78% (7/9 patients) gliotic response after a range of 7-262 months following DBS implantation is lower than the overall rate of gliotic responses reported in the literature (92%, 22/24 patients). Notably, one of our patients without gliosis had DBS for 7 months. Previous studies have attributed the reaction of the brain parenchyma to the electrode itself and/or the chronic stimulation.^32^ More study is needed to understand if these factors, such as the biocompatibility of the implanted electrodes, the variability in the stimulation parameters, degradation of the metal contacts, and the length of stimulation (and others), could be implicated in the process of gliotic scar formation. The clinical implication (effect upon efficacy variation) of the gliotic response around the DBS electrode is poorly described. The plausibility that an increased gliotic response negatively affects DBS efficacy seems reasonable and intuitive. However, in our study, from the 7 patients who developed gliotic changes, 5 had a 32% reduction in their LEDD at the last follow-up compared to pre-DBS dosage. This contrasted with the 2 patients who needed a greater than 50% LEDD increase over their post-implantation course. Of note, these patients (4 and 6 from Table 1) had the lowest pre-DBS LEDD levels and a prolonged DBS treatment (65 and 94 months, respectively). Also, patient 6 was the only one who received stimulation of the globus pallidus interna, which is known to not reduce LEDD after DBS compared to patients treated with STN DBS. Given the variability in our findings, the limited sample size, and the lack of standardization in the assessment of gliosis around the DBS electrode, additional studies will be needed with objective clinical findings, such as LEDD and TEED, compared to in depth histological evaluation, such as volume and density of gliosis, before robust conclusions can be made. Separating the gliosis produced by the implantation lesion and the simulation-induced effects is also relevant for optimizing clinical efficacy. Thus, we recommend that future studies evaluate the potential correlation between the gliotic scar and the clinical efficacy of DBS treatment to provide needed insights for developing biocompatible materials, optimizing stimulation parameters, and implantation techniques that optimize patient outcomes.

### Additional neurohistopathologic findings

Evidence of additional markers of local tissue response (to the implanted electrodes), such as activated microglia, reactive astrocytosis, giant multinuclear cells, hemosiderin-laden macrophages, and neuronal loss, have been reported in previous studies.^2,4,8^ In our series, only one (11%) case (patient 2) demonstrated activated microglia infiltration, which was lower than the 53% we found in the literature review. The presence of activated microglia lacks specificity but supports the presence of an ongoing inflammatory response regardless of the chronicity of the electrode implantation. Furthermore, in the literature review, we found additional tissue responses such as astrogliosis (53%), multinucleated giant cells (32%), and neuronal loss (37%). Although these findings are unspecific and were not assessed in our series, their description, characterization, and role in the therapeutic action of DBS are of great interest. Support for more postmortem neurohistopathologic after DBS is also supported by our findings of change in diagnosis over time in our series and the literature. In our study 1 patient was confirmed to have MSA after DBS implantation for PD, while in the literature, 2 cases^18,30^ had a similar update to MSA after postmortem assessment. The lack of consistent efficacy in these cases confirms the need for an accurate diagnosis to obtain the best clinical results. Clinical research would benefit from pathologically confirmed diagnosis to ensure data is accurate. Additional knowledge about DBS and its mechanisms could allow for the inclusion of other indications in which the therapy could have potential benefits.

### Future directions

Although the neurohistopathological changes observed in the tissue surrounding the DBS electrodes show characteristics of a classical foreign body reaction, the presence of additional changes (around the active electrodes) and their underlying mechanisms are yet to be elucidated. The response of different cellular lines (neurons, glial cells, immune cells, and vasculature-related cells) to electrical stimulation is not established.^1^ Determination of the influence of electrical currents (at varying intensities and charge densities) on neuronal signaling, neural function, astrocyte response, and overall cellular metabolism could provide valuable insights to tailor and optimize the stimulation parameters to favor minimal injury, maximal long-lasting efficacy, and expansion of the contemporary indications for DBS. Additionally, the development and testing of electrodes (and coatings) with novel functional biomaterials aiming to minimize the immune response, decrease tissue resistance to stimulation (impedance), favor the attraction of the neuronal processes (potentially enhancing the benefits of DBS), and optimize the charge transfer density. Such efforts are underway employing nanomaterials and nanoparticles that minimize the tissue reaction, favor neural recordings, and improve charge transfer efficiency.^46^ Despite this, the use of postmortem neurohistopathological data continues to provide valuable and fundamental insights to guide future directions. However, the postmortem literature is largely dominated by studies with low-level evidence (case reports and case series), lacking standard tissue handling and assessment protocols. The formation of an integrated network of researchers, the adoption of common protocols for the collection and management of specimens, and the implementation of new techniques for tissue evaluation seem to be the natural next step in the evolution of the study of human neurodegenerative and neuropsychiatric disease and the role of DBS with them.^47,48^

### Limitations

We acknowledge that our study has limitations. First, the sample size was small, and the analysis was retrospective. Next, the clinical outcome data were limited by availability and patient follow-up Additionally, the handling, processing, and evaluation of the neurohistopathological samples were performed according to local protocols, which limits the comparison and generalizability of our findings. Regarding the systematic review, the included studies were small (case reports and series) and retrospective. The clinical data and postmortem neurohistopathological findings were heterogeneously reported based on the individual selection and preference of the authors from each study. However, considering the limited availability of such data and the lack of standardization on the protocols for neurohistopathological tissue handling, we included all the available reports aiming to comprehensively summarize the existing literature.

## CONCLUSION

Our findings confirm the presence of a local tissue reaction, including gliosis and activated microglial infiltration around the implanted DBS electrodes. The presence of additional histologic changes has been reported in the literature however, the implication of these findings needs to be clearly established. The effect of the local changes on the clinical efficacy of DBS is not clearly established. As the continual effort to expand the use, efficacy, and safety of DBS for neuromodulation and neuroaugmentation moves forward, the value of postmortem neurohistopathological evidence will undoubtedly shed more light on the mechanism behind this powerful clinical and surgical tool. Further DBS postmortem studies and standardization of tissue processing are needed to expand the understanding of the interaction between the local brain parenchyma and the implanted DBS electrode.

## Data Availability

All data produced in the present study are available upon reasonable request to the authors.

## ACKNOWLEDGMENTS

The authors would like to acknowledge the incredible contribution of the patient donors and their families, as this research would not be possible without their generous gift to the advancement of science. They would also like to acknowledge the support staff at the University of Iowa who worked tirelessly behind the scenes to keep our facilities running. If there are additional comments/suggestions, please let us know.

## ABBREVIATIONS

DBS: deep brain stimulation
LEDD: Levodopa equivalent daily dose
MSA: multiple system atrophy
PD: Parkinson’s disease
TEED: total electrical energy delivered.

