## Supplemental tables for "Neurohistopathological Findings of the Brain Parenchyma After Long-Term Deep Brain Stimulation: Case Series and Systematic Literature Review"

**SUPPLEMENTAL MATERIAL**

**Table of Contents**

**Supplemental material Table 1.** Search strategies for PubMed (MEDLINE)	2

**Supplemental material Table 2.** Characteristics of the medical management and stimulation parameters of the patients who underwent DBS implantation	3

**Supplemental material Table 3.** Characteristics of the postmortem neurohistopathological findings around the DBS electrode track for epilepsy	4

**Supplemental material Table 4.** Characteristics of the postmortem neurohistopathological findings around the DBS electrode track for pain	5

**Supplemental material Table 5.** Characteristics of the postmortem neurohistopathological findings around the DBS electrode track for other conditions	6

**References**	7

**Table 1**. Search strategy for Pubmed (MEDLINE) *

| **Database** | **Search query** | **Results** |
| --- | --- | --- |
| PubMed | (DBS [tiab] OR "deep brain stimulation" [tiab] OR "high frequency stimulation" [tiab]) AND ("post-mortem" OR "pathologic" OR "pathology" OR "neuropathology" OR "clinicopathologic" OR "postmortem" OR "autopsy" OR "clinicopathological" OR "fatal outcome") | 1249 |
| * End-of-search date: August 9, 2023 | | |

**Table 2.** Characteristics of the medical management and stimulation parameters of the patients who underwent DBS implantation

| **DBS case** | **DBS target** | **Pre-DBS implantation** | **6 (± 3) month post-DBS** | | | | **Last available follow-up** | | | | **Duration of DBS (months)** |
| --- | --- | --- | --- | --- | --- | --- | --- | --- | --- | --- | --- |
|  |  | **LEDD (mg)** | **LEDD (mg)** | **TEED (J⋅s)** | | | **LEDD (mg)** | **TEED (J⋅s)** | | |  |
|  |  |  |  | **Left-sided electrode** | **Right-sided electrode** | **Combined** |  | **Left-sided electrode** | **Right-sided electrode** | **Combined** |  |
| 1 | BL STN | 1,500 | 1,400 | 3 | 3 | 6 | 1,300 | 85 | 77 | 162 | 24 |
| 2 | BL STN | 1,000 | NA | NA | NA | NA | 900 | NA | NA | NA | 140 |
| 3 | BL STN | 1,100 | 1,100 | 8 | 34 | 95 | 750 | 61 | 160 | 316 | 126 |
|  | L VIM |  |  | 53 | - |  |  | 95 | - |  |  |
| 4 | BL STN | 700 | NA | NA | NA | NA | 1,170 | NA | NA | NA | 65 |
| 5 | BL STN | 925 | 750 | 73 | 68 | 141 | 500 | 116 | 212 | 328 | 262 |
| 6 | BL GPi | 840 | 715 | 70 | 96 | 166 | 2,625 | 157 | 157 | 313 | 94 |
| 7 | BL STN | 1,820 | 1,820 | 5 | 16 | 21 | 1,410 | 23 | 44 | 67 | 7 |
| 8 | BL STN | 1,500 | 1,100 | 23 | 24 | 47 | 1,300 | 28 | 18 | 46 | 13 |
| 9 | BL STN | 1,936 | 1,390 | 20 | 47 | 67 | 850 | 15 | 31 | 46 | 62 |
| **Median [IQR]** |  | 1,100  [883 – 1,660] | 1,100  [750 – 1,400] | 22  [6 – 66] | 34  [16 – 68] | 67  [21 – 141] | 1,170  [800 – 1,355] | 73  [24 – 111] | 77  [31 – 160] | 162  [46 – 316] | 65  [19 – 133] |

Abbreviations: BL, bilateral; DBS, deep brain stimulation; GPi, globus pallidus interna; IQR, interquartile range; L, left; LEDD, Levodopa equivalent daily dose; NA, not available; STN, subthalamic nucleus; TEED, total electrical energy delivered; VIM, ventralis intermedius thalamic nucleus.

**Table 3.** Characteristics of the postmortem neurohistopathological findings around the DBS electrode track for epilepsy

| **Author, year** | **DBS case n.** | **Age at DBS** | **Sex** | **DBS targets** | **Electrode surface material** | **Clinical efficacy after DBS** | **Clinical**  **decline after DBS** | **Duration in months** | **Cause of death** | **Gliosis at DBS site** | **Activated microglia at DBS lead** | **Neuronal loss at DBS site** | **Other findings at DBS site** |
| --- | --- | --- | --- | --- | --- | --- | --- | --- | --- | --- | --- | --- | --- |
| Robertson et al., 1979[1] | 1 | NR | M | Cerebellum: BL anterior lobe and rostral portion of posterior lobe | Platinum | Yes | No | 15 | Seizure | Yes | Yes | Yes | Thick capsule around electrode |
| “ | 2 | NR | F | Cerebellum: BL anterior lobe and rostral portion of posterior lobe | Platinum | No | No | 8 | Drowning | Yes | Yes | Yes | Giant cells |
| “ | 3 | NR | M | Cerebellum: BL anterior lobe and rostral portion of posterior lobe | Platinum | No | No | 6.6 | Suicide | Yes | Yes | Yes | - |
| Wright and Weller, 1983[2] | 4 | 21-25 | M | Cerebellum: BL upper surface | NR | Yes | No | 16 | Seizure | Yes | No | Yes | No inflammatory changes |
| Pilitsis et al., 2008[3] | 5 | 21-25 | M | BL anterior nucleus of the thalamus | Platinum/Iridium | No | No | 8 | SUDEP | Yes | Yes | Yes (minimal) | Activated microphages |
| Giordano et al., 2020[4] | 6 | 6-10 | M | BL globus palidus internus | Platinum/Iridium | Yes | No | 2 | Infection | Yes | Yes | Yes (minimal) | Lymphocyte infiltration with giant multinucleated cells |

Abbreviations: BL, bilateral; DBS, deep brain stimulation; F, female; M, male; NR, not reported; n., number; SUDEP, sudden unexpected death in epilepsy.

**Table 4.** Characteristics of the postmortem neurohistopathological findings around the DBS electrode track for pain

| **Author, year** | **DBS case n.** | **Age at DBS** | **Sex** | **Pain related condition** | **DBS targets** | **Electrode surface material** | **Pain releif after DBS** | **Worse pain after DBS** | **Duration in months** | **Cause of death** | **Gliosis at DBS site** | **Activated microglia at DBS lead** | **Neuronal loss** | **Other findings at DBS site** |
| --- | --- | --- | --- | --- | --- | --- | --- | --- | --- | --- | --- | --- | --- | --- |
| Hosobuchi et al., 1977[5] | 1 | 51-55 | M | Rectum Ca | Left central gray | Platinum/Iridium | Yes | No | 5 | NR | NR | NR | NR | - |
| “ | 2 | 56-60 | M | Diabetic neuropathy | Left central gray | Platinum/Iridium | Yes | No | 7 | NR | NR | NR | NR | - |
| Gybels et al., 1980[6] | 3 | NR | NR | Head Ca (pituitary) | Periventricular and periaqueductal gray | Stainless steel | Yes | No | 6 | NR | Yes | Yes | Yes | Necrosis and Gitter cells |
| “ | 4 | NR | NR | Head Ca (nasal cavity) | Periventricular and periaqueductal gray | Platinum/Iridium | No | No | 17 | NR | Yes | Yes | Yes | Spongiosis |
| “ | 5 | NR | NR | Head Ca (tongue and maxilla) | Periventricular and periaqueductal gray | Platinum/Iridium | No | No | 9 | NR | Yes | Yes | Yes | Spongiosis |
| “ | 6 | NR | NR | Breast Ca metastasis (hip) | Periventricular and periaqueductal gray | Platinum | No | No | 0.5 | NR | No | No | No | Giant cells and necrosis |
| “ | 7 | NR | NR | Head Ca (maxillary sinus) | Periventricular and periaqueductal gray | Platinum | No | No | 0.75 | NR | No | Yes | No | Necrosis and Gitter cells |
| Boivie and Meyerson, 1982[7] | 8 | 66-70 | F | Rectal Ca metastasis (multiple) | Right thalamus | Platinum/Iridium | Yes | No | 6 | Malignancy | Yes | Yes | Yes | - |
| “ | 9 | 56-60 | F | Rectal Ca metastasis (pelvis) | Right thalamus | Platinum/Iridium | Yes | No | 4 | Malignancy | Yes | Yes | Yes | Gitter cells |
| “ | 10 | 41-45 | M | Lung Ca metastasis (spine) | BL thalamus | Platinum/Iridium | Yes | No | 3 weeks | Malignancy | No | Yes | No | - |
| “ | 11 | 51-55 | F | Bladder Ca metastasis (spine) | Right thalamus | Platinum/Iridium | No | No | 12 | Malignancy | Yes | Yes | Yes | - |
| “ | 12 | 51-55 | M | Rectal Ca metastasis (multiple) | Right thalamus (x2) | Platinum/Iridium | Yes | Yes | 17 | Malignancy | Yes | Yes | Yes | - |
| Baskin et al., 1986[8] | 13 | 36-40 | M | Laryngeal and esophageal Ca | Left periaqueductal gray | Platinum | Yes | Yes | 3 | Malignancy | No | NR | NR | - |
| “ | 14 | 51-55 | M | Rectal Ca | BL periaqueductal gray | Platinum | Yes | Yes | 5 | Malignancy | No | NR | NR | - |
| “ | 15 | 56-60 | M | Diabetic neuropathy | Left periaqueductal gray | Platinum | Yes | No | 7 | Diabetes | No | NR | NR | - |
| “ | 16 | 61-65 | F | Rectal Ca | BL periaqueductal gray | Platinum | Yes | No | 1 | Malignancy | No | NR | NR | - |
| “ | 17 | 61-65 | M | Colon Ca | Right periaqueductal gray | Platinum | Yes | No | 2 | Malignancy | No | NR | NR | - |
| “ | 18 | 61-65 | M | Lung Ca | Right periaqueductal gray | Platinum | Yes | Yes | 2 | Malignancy | No | NR | NR | - |
| “ | 19 | 41-45 | F | Breast Ca metastasis | Right periaqueductal gray | Platinum | Yes | No | 2 | Malignancy | No | NR | NR | - |
| Kuroda et al., 1991[9] | 20 | 71-75 | M | Bilateral neuropathic pain | Right thalamus | NR | Yes | Yes | 20 | Tuberculous pleuritis | Yes | NR | Yes | - |

Abbreviations: BL, bilateral; Ca, cancer; DBS, deep brain stimulation; F, female; M, male; n., number; NR, not reported.

**Table 5.** Characteristics of the postmortem neurohistopathological findings around the DBS electrode track for other conditions

| **Author, year** | **DBS case n.** | **Indication** | **Age at DBS** | **Sex** | **DBS targets** | **Electrode surface material** | **Clinical efficacy after DBS** | **Clinical decline after DBS** | **Duration in months** | **Cause of death** | **Gliosis at DBS site** | **Activated microglia at DBS lead** | **Neuronal loss** | **Other findings at DBS site** |
| --- | --- | --- | --- | --- | --- | --- | --- | --- | --- | --- | --- | --- | --- | --- |
| Boockvar et al., 2000[10] | 1 | BL essential tremor | 46-50 | F | BL ventralis intermedius nucleus | Platinum/Iridium | Yes | No | 16 | Rhinological surgery complication | Yes | NR | No | Iron deposits within tracks |
| Burbaud et al., 2002[11] | 2 | Chorea-acanthocytosis | 41-45 | M | BL ventral oral of motor thalamus | Platinum/Iridium | Yes | No | 24 | Unknown cause | Yes | NR | No | Right track: perivascular lymphocytic infiltrates and iron deposits |
| Gross et al., 2004[12] | 3 | BL action tremor | 61-65 | M | Right ventralis intermedius nucleus | Platinum | Yes | No | 12 | Cryptogenic cirrhosis | Yes | NR | NR | 3 additional microelectrode tracks |
| Valldeoriola et al., 2006[13] | 4 | Diffuse Lewy body disease * | 71-75 | M | BL subthalamic nuceus | Platinum/Iridium | Yes | No | 42 | Bronchopneumonia | Yes | NR | NR | Infiltrates of T lymphocytes ** |
| McClelland et al., 2007[14] | 5 | Diffuse Lewy body disease * | 66-70 | M | BL subthalamic nuceus | Platinum/Iridium | Yes | No | 40 | Progressive dementia | Yes | NR | No | - *** |
| DiLorenzo et al., 2010[15] | 6 | BL essential tremor | 71-75 | F | BL ventralis intermedius nucleus | Platinum | Yes | No | 146 | Non-pathologic | Yes | NR | No | Macrophages, multinucleated giant cells, and T lymphocytes |
| Hughes et al., 2011[16] | 7 | Dystonic tremor | 51-55 | M | BL ventralis intermedius nucleus | Platinum/Iridium | Yes | No | 48 | Viral myocarditis | Yes | Yes | Yes | Perivascular lymphocytes |

* Patient was initially diagnosed with Parkinson’s disease. ** Lewy bodies and Lewy neurites in the substantia nigra pars compacta, locus coeruleus, raphe nuclei, dorsal nucleus of the vagus, and hypoglossal nerve. *** Lewy body-containing neurons and neurites in the dorsal nucleus of the vagus, nucleus, caeruleus, pars compacta of the substantia nigra, hypothalamus, substantia innominata, and throughout the neocortex.

Abbreviations: BL, bilateral; DBS, deep brain stimulation; F, female; M, male; n., number; NR, not reported.
